# 50 Hz cortical stimulation increases interictal epileptiform discharges at the seizure onset zone

**DOI:** 10.1101/2025.03.03.25322984

**Authors:** Ahdyie Ahmadi, Hellen Kreinter Rosembaun, Benjamin Corrigan, Mohamad Abbass, Greydon Gilmore, Nasim Mortazavi, Giovanni Pellegrino, Jorge G. Burneo, David A. Steven, Jonathan C. Lau, Keith W. MacDougall, Michelle-Lee Jones, Julio Martinez-Trujillo, Ana Suller Marti

## Abstract

More than 15 million patients worldwide suffer from drug-resistant epilepsy (DRE). Surgical removal of the seizure onset zone (SOZ)—the brain region(s) from where seizures arise—is the best available treatment for these patients, with post-surgical outcomes depending on the successful identification and resection of the SOZ. Most commonly, SOZ mapping localizes ictal activity occurring spontaneously or evoked by cortical stimulation (CS) during presurgical evaluation of patients with epilepsy using stereoelectroncephalography (SEEG). Mapping events such as interictal epileptiform discharges (IEDs), paroxysmal hypersynchronic electrical discharges that often occur outside ictal discharges or during CS, have been less used for SOZ localization.

We test the hypothesis that IEDs triggered by CS via SEEG investigation can contribute to the mapping of the SOZ. We evaluated the impact of CS on IEDs in patients investigated with SEEG on epilepsy surgery investigation. We recorded intracranial signals from 30 DRE patients (seizure-free post-surgery). Bipolar and high frequency (50 Hz) CS was performed with a pulse width of 300 µs and current spanning 1–8 mA. IEDs were automatically detected pre- and post- stimulation, and their normalized absolute changes were quantified within and outside the SOZ (identified by ictal discharges).

We found that IED rates significantly increased post-stimulation compared to pre-stimulation within the SOZ, while no significant change was observed outside the SOZ (Linear mixed effect model, p-value <0.001, and AUC=98% for SOZ and 71% for non-SOZ). This effect was present regardless of whether the stimulation was applied to the SOZ or non-SOZ regions, indicating a broader effect of stimulation on the SOZ.

Our results offer a quantitative tool for identifying epileptogenic areas in patients with DRE, enhancing the mapping and localization of the SOZ and potentially improving surgical outcomes.

## Introduction

Epilepsy is a chronic neurological disorder affecting about 50 million people worldwide ^1^, with over 15 million experiencing drug-resistant epilepsy (DRE) despite appropriate medication trials.^2,3^ Around 60% of DRE cases involve focal epilepsy, where seizures originate from a specific brain region called the seizure onset zone (SOZ).^4^ Surgical resection or neuromodulation can eliminate seizures if the SOZ is accurately identified through a thorough evaluation.^5^ When non-invasive tests are inconclusive, invasive monitoring using stereoelectroencephalography (SEEG) and electrocorticography (ECoG), gold-standard techniques for SOZ localization, is performed by implanting electrodes to record seizure activity and guide treatment.^6,7^

However, seizures are unpredictable and often infrequent, making ictal recordings costly, time-consuming, and risky.^8^ Furthermore, even with intracranial recordings and qualitative analyses to define the SOZ, predicting surgical outcomes for individual patients remains difficult, with success rates ranging from 30% to 70%.^2^ To address this, a quantitative approach that estimates the most epileptogenic regions through the identification of relevant biomarkers can be proposed .^9^ Intracranial recordings from patients with epilepsy frequently capture other electrophysiological disturbances, such as interictal epileptiform discharges (IEDs), a type of paroxysmal electrical event generated by the synchronous activity of a population of neurons in epileptic brains.^10^ IEDs occur more often than seizures and can expedite and enhance the identification of the SOZ.^11^ At least one study has shown that ictal activity can predict the occurrence of IEDs, suggesting that these events are linked to activation of epileptogenic networks.^12^ Thus, it is possible that activating epileptogenic networks within the SOZ lead to increase in the rate of IEDs within the zone and can therefore be used as a biomarker to plan surgical resections.

As part of the presurgical evaluation, extraoperative electrical stimulation (CS) at a frequency of 50 Hz is primarily used to map the eloquent cortex, helping to prevent or minimize the risk of unintended neurological damage.^13^ It was also documented that cortical stimulation-induced seizures are associated with the surgical outcome.^14^ Some studies have reported the effects of intraoperative CS on IEDs detected in intracranial recordings using subdural grids. Nakatani et al. found that high-frequency stimulation of SOZ decreases the amplitude of spikes and the number of IEDs.^15^ Similar results were documented by other studies.^16,17^ However, SEEG 50Hz CS have been utilized to trigger seizures. Currently, little is known about how IEDs rates change in response to 50 Hz cortical stimulation and whether it could provide insights into localization of the SOZ. In this study, we investigate this issue.

We hypothesize that high-frequency (50 Hz) intracortical stimulation increases the number of evoked IEDs, particularly at the SOZ region defined by mapping of ictal activity. Cortical stimulation mimics seizure and at least one study has suggested that IEDs increase after seizures.^18^ In this study, we aim to evaluate changes in IEDs before and after stimulation, depending on whether the stimulated region is within or outside the SOZ. Our ultimate goal is to assess whether this measurement can aid in localizing the SOZ.

## Materials and methods

### Patients

We recruited patients with drug-resistant epilepsy (18 years and older) who underwent invasive presurgical evaluation (phase II) at our centre. The study received approval from our institution’s research ethics board, and all participants provided written informed consent before they were included in this study. Our dataset comprised 135 patients at the time of analysis, of which 108 underwent high-frequency cortical stimulation (50Hz). Patients who received any form of neuromodulation, including deep brain stimulation (DBS), responsive neurostimulation (RNS), or vagus nerve stimulation (VNS), were excluded from the analysis. The final cohort included 26 patients who underwent resection and four patients who underwent ablation, all of whom became seizure-free following intervention, with a minimum of 6 months of follow-up (**Fig. 1**). This selection ensured that the treated areas included the SOZ. A total of 30 patients (18 female; age range: 18-59 years; mean age = 31.36 years; standard deviation = 11.12) were analyzed in this study. Detailed patient information is summarized in **Table 1**.

**Figure 1.**
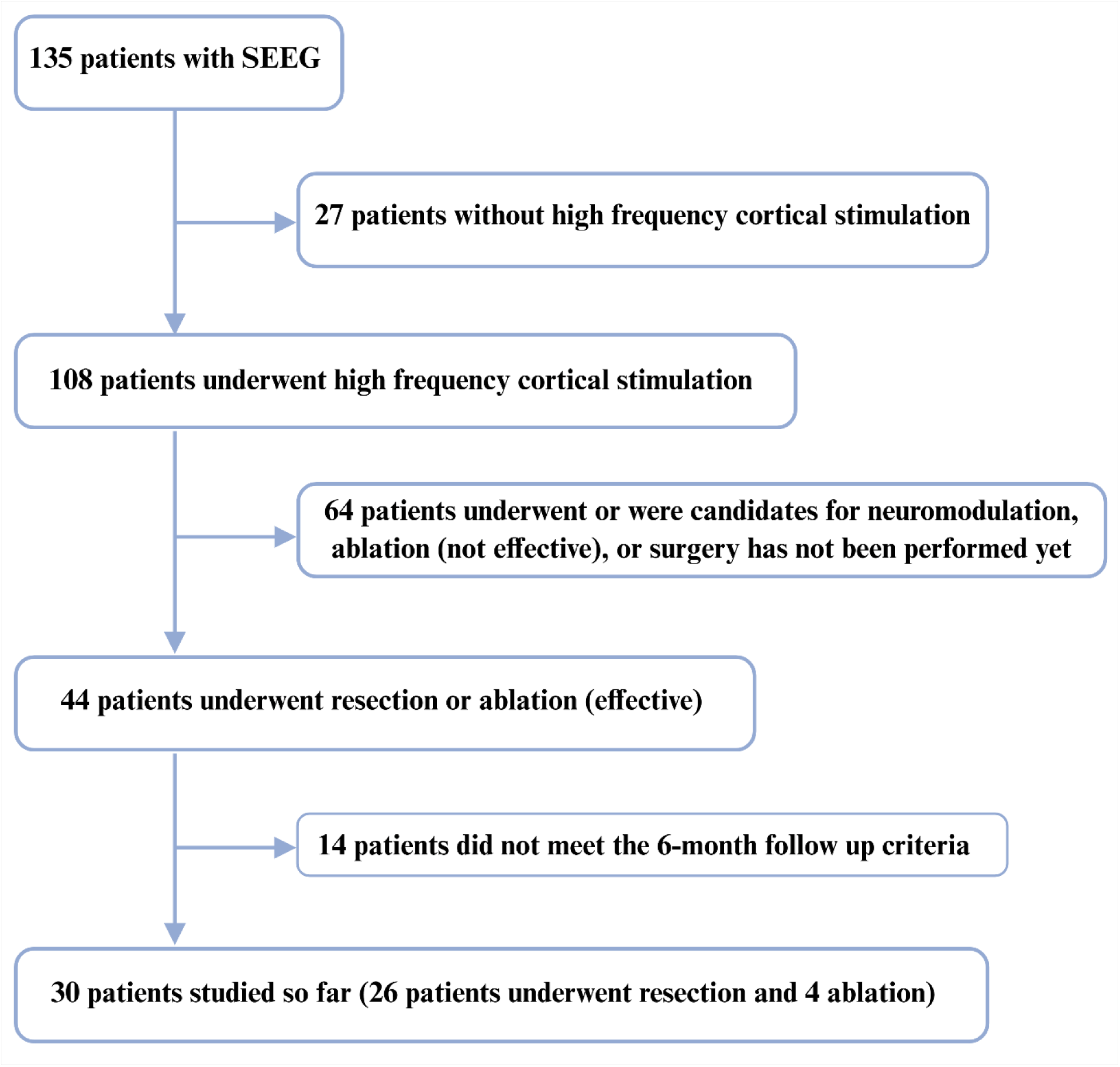
Patients’ flowchart. Patients included in the study underwent high-frequency cortical stimulation without any neuromodulation. Only patients who underwent ablation or resection, became seizure-free, and completed the 6-month follow-up were included in the final analysis. SEEG: Stereoelectroencephalography. (Created in BioRender. Ahmadi, A. (2025) https://BioRender.com/4uhf92z)

**Tables.**

| Number | Number of electrodes | Number of stimulation trials | Diagnosis | Pathology | Intervention | Maximum delivered current | Total SOZ contacts |
| --- | --- | --- | --- | --- | --- | --- | --- |
| 1 | 10 | 11 | Lt TLE | HS | R | 6 | 17 |
| 2 | 8 | 17 | Rt MTLE | NS | R | 5 | 14 |
| 3 | 13 | 26 | Lt TLE | RG | R | 8 | 22 |
| 4 | 8 | 42 | Lt MTLE | RG | R | 6 | 14 |
| 5 | 11 | 36 | Lt TLE | NO | R | 6 | 19 |
| 6 | 9 | 21 | Lt OLE/TLE | RG | R | 8 | 2 |
| 7 | 10 | 59 | Lt FLE/ TLE | MTS | R | 10 | 17 |
| 8 | 12 | 8 | Lt TLE | NO | R | 7 | 21 |
| 9 | 7 | 11 | Lt MTLE | NO | R | 10 | 2 |
| 10 | 11 | 8 | Lt TLE | MTS | R | 10 | 19 |
| 11 | 12 | 29 | Rt FLE | FCD | R | 8 | 21 |
| 12 | 19 | 11 | Rt MFLE | NO | R | 6 | 44 |
| 13 | 8 | 24 | Rt FLE | FCD | R | 1 | 14 |
| 14 | 9 | 2 | Rt MTLE | MTS | R | 3 | 2 |
| 15 | 10 | 1 | Rt TLE | RG | R | 1 | 20 |
| 16 | 14 | 10 | Rt TLE | NO | R | 6 | 22 |
| 17 | 8 | 40 | Rt TLE | HS | R | 10 | 14 |
| 18 | 13 | 64 | Lt FLE | NO | R | 10 | 22 |
| 19 | 10 | 50 | Rt TLE | RG | R | 10 | 17 |
| 20 | 14 | 24 | Rt TLE | NS | R | 8 | 22 |
| 21 | 9 | 28 | Rt FLE | NO | A | 5 | 16 |
| 22 | 16 | 24 | Lt ILE | NO | A | 8 | 28 |
| 23 | 13 | 36 | Lt OLE | FCD | A | 8 | 22 |
| 24 | 8 | 6 | Rt FLE | NS | R | 1 | 14 |
| 25 | 8 | 10 | Lt MTLE | NO | R | 6 | 14 |
| 26 | 8 | 31 | Rt MTLE | MTS | R | 5 | 14 |
| 27 | 15 | 21 | Rt FLE | NS | R | 6 | 22 |
| 28 | 14 | 24 | Lt MTLE | MTS | R | 6 | 22 |
| 29 | 9 | 23 | Lt MTLE | MTS | R | 6 | 21 |
| 30 | 12 | 12 | Rt TLE | MTS | A | 6 | 20 |

Lt: Left, Rt: Right, TLE: Temporal lobe epilepsy, MTLE: Mesial temporal lobe epilepsy, OLE: Occipital lobe epilepsy, FLE: Frontal lobe epilepsy, HS: Hippocampal sclerosis, RG: Reactive gliosis, MTS: Mesial temporal sclerosis, ILE: Insular lobe epilepsy, FCD: Focal cortical dysplasia, NS: Nonspecific changes, NO: No information, A: Ablation, R: Resection, Number of electrodes: Number of SEEG shafts, SOZ: Seizure onset zone,

### Electrode implantation

SEEG is an invasive technique for monitoring brain electrical activity utilizing depth electrodes. Depth electrodes typically feature a diameter of 0.86 mm with platinum contacts, each 2.29 mm long (Ad Tech Medical®, WI, USA). The number of contacts per electrode ranges from four to 18, with an intercontact distance of 3 to 6 millimetres, based on the electrode type. Before surgery, the placement of electrodes was planned to target specific areas of the brain potentially involved in seizure initiation. Following implantation, the position of electrodes was confirmed using computed tomography co-registered with preoperative MRI.^13^

### Electrical Cortical Stimulation

Electrical cortical stimulation was conducted following the recording of interictal data and typical electroclinical seizures. During stimulation, a precise 3-dimensional map of the contacts in relationship to cortical areas (topography) and other brain structures was readily available. Before the CS session, we established a list of channels to be stimulated. The channels were categorized into the most epileptogenic and least epileptogenic areas based on spontaneous IED activity and ictal discharges observed during presurgical monitoring and typical electroclinical seizures. The least epileptogenic areas are characterized as regions that display minimal or no IEDs during monitoring and show no significant involvement in ictal discharges during the spontaneous seizure evolution and propagation. These areas were considered less likely to contribute to spontaneous seizure generation. On the other hand, the more epileptogenic areas are defined as regions exhibiting the highest frequency of spontaneous IEDs during presurgical monitoring, as well as consistent involvement in ictal discharges during spontaneous seizures. These areas were typically associated with the SOZ and are primary targets for CS or surgical resection. Contacts covering the putative SOZ were reserved for the end of the stimulation session, due to the potential occurrence of seizures.^13^ The adjacent contact pairs selected for stimulation were determined by the clinical team based on the implantation scheme and the clinical hypothesis regarding the epileptogenic network. Stimulation was delivered through pairs located both within the suspected SOZ and outside of it, to assess the network-wide effects of cortical stimulation.

Electrical stimuli were delivered using the Nicolet Cortical Stimulation device (Natus)®. Stimulation parameters typically used range from 1 to 8 mA, with a pulse width of 300 μs (the selection of this pulse width was based on the standard protocols in our center, noting that stimulation parameters may vary across modalities such as thalamic, responsive, or VNS), at a frequency of 50 Hz (in regions with 50 Hz alternative current (AC), a different frequency may be used). For this analysis, we did not include low-frequency stimulation (see supplementary **Table 1**).

Stimulation was applied at lower intensities for 1–5 seconds or until clinical symptoms, provoked seizures or after-discharges, appear. After-discharges are characterized by rhythmic waves, repetitive epileptiform potentials, or a combination of both, triggered by the stimulus .^19^ These parameters were used to stimulate both the non-SOZ and SOZ (**Fig. 2A**). Stimulation began at 1 mA, and intensity was incrementally increased until clinical symptoms appeared, after-discharges or seizures were triggered, or the maximum intensity was reached. The total duration of stimulation and the delivered charge were kept within established safety limits.

**Figure 2.**
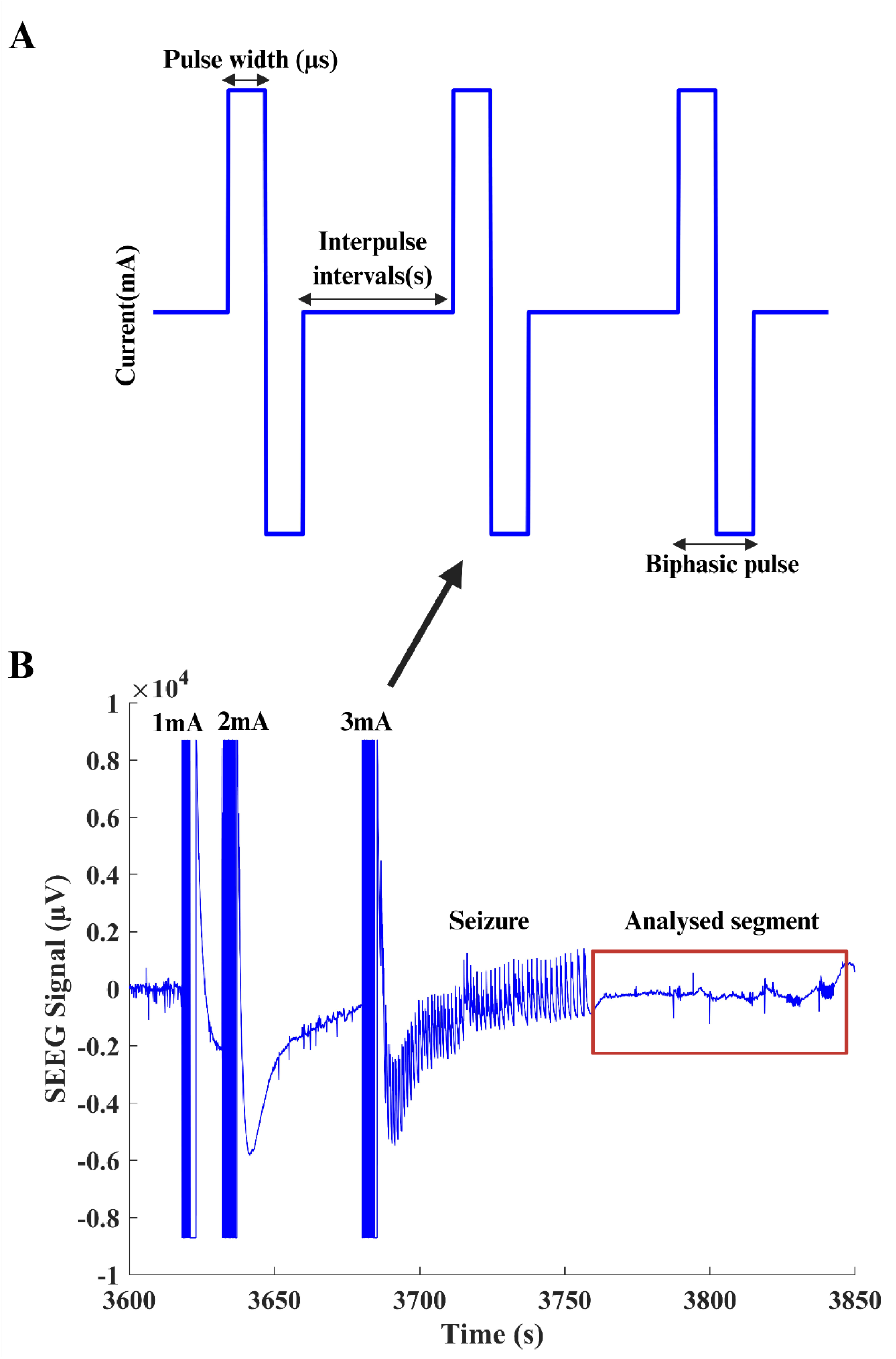
Analysed segment of the signal. (**A)** Schematic representation of the biphasic stimulation mode. **Current (mA):** The intensity of the electrical stimulation delivered in each pulse, measured in milliamperes (mA). **Pulse width (µs):** The duration of each stimulation pulse, measured in microseconds (µs). **Interpulse interval (s):** The time between consecutive stimulation pulses, measured in seconds (s). **Biphasic pulse:** A stimulation pulse that has two phases of opposite polarity, typically one positive followed by one negative phase, to reduce net charge delivery and tissue damage. (**B)** SEEG signal during stimulation pulses. The x-axis displays time during the session in seconds and the y-axis the SEEG signal amplitude. Three stimulation trials are shown with amplitudes of 1 mA, 2 mA, and 3 mA. IEDs were extracted following the termination of stimulation trial and clinical symptoms, a seizure in this case (red rectangle). SEEG: Stereoelectroencephalography. Panels A and B were generated in MATLAB and combined using Biorender. (Created in BioRender. Ahmadi, A. (2025) https://BioRender.com/onphgyx)

### Data Preprocessing

Intracranial recordings were sampled at 2048 Hz without applying a digital filter, and were collected at three stages: before, during, and after stimulation. Before CS, electrode locations were verified through computed tomography (CT) scans. Noisy channels were excluded to ensure optimal data quality. For referencing, we used a bipolar derivation technique, subtracting consecutive channel signals, as stimulation was delivered in bipolar mode. The data was segmented into pre-stimulation and post-stimulation periods. For each pair of channels, a pre-stimulation (baseline) epoch was defined using the initial minutes of the recording at the beginning of the stimulation session, before any stimulation was delivered. This epoch was free of stimulation artifacts, seizures, and after-discharges, ensuring that it accurately reflected the unperturbed state of the brain. Each post-stimulation trial for a given pair of channels was then compared to the corresponding baseline epoch from the same pair of channels. Post-stimulation period was defined as the signal segment following the application of maximum current over a pair of channels, as illustrated in **Fig. 2B**. Additionally, Notch biquad filters with a 4 Hz bandwidth were used to attenuate 60 Hz hum noise. Visual inspection was conducted to identify and remove potential artifacts. The average length of the analyzed segment, before and after stimulation, was 5 minutes. From the cleaned signals, provoked IEDs were extracted. The preprocessing and IED extraction steps are detailed in **Fig. 3**. Electrode contacts located in the white matter were excluded from the analyses, as they are not expected to generate or reliably record interictal discharges. Only contacts localized in gray matter structures were included in both stimulated and non-stimulated contact analyses.

**Figure 3.**
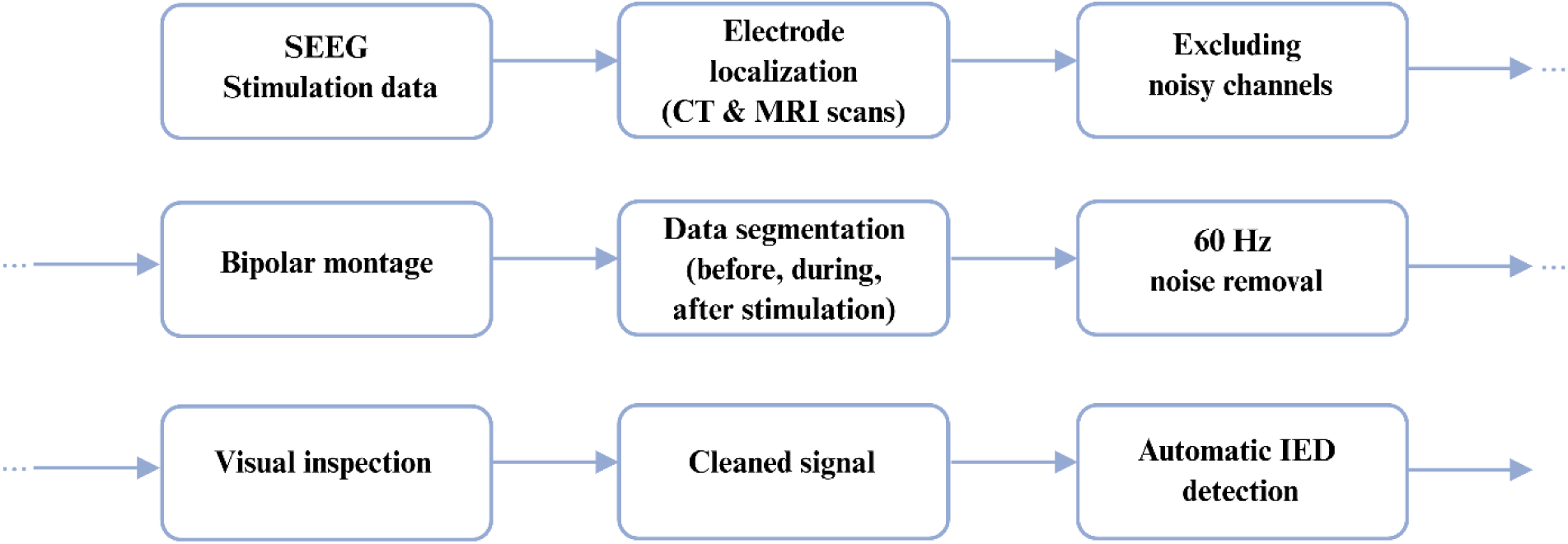
Preprocessing Pipeline. Overview of the preprocessing steps applied to SEEG stimulation data, including electrode localization, noise and artifact removal, montage application, data segmentation, and automatic IED detection. SEEG: Stereoelectroencephalography; IED: Interictal Epileptiform Discharge; CT: Computed Tomography; MRI: Magnetic Resonance Imaging. (Created in BioRender. Ahmadi, A. (2025) https://BioRender.com/dmljse4)

### IED extraction

IED detection was performed using the algorithm developed by Janca et al.^20^, which adaptively models the statistical distribution of signal envelopes to distinguish IEDs from background activity. The algorithm detects IEDs based on local increases in signal energy, particularly within the 14.3–50 Hz frequency range, and has a reported sensitivity of 97.4 ± 12.2%. In our implementation, signals were filtered between 10–60 Hz using zero-phase 8th-order Chebyshev Type II digital filters in MATLAB Rb2022. This method is also capable of identifying low-voltage IEDs that are often overlooked during visual inspection and may offer additional clinical insights.

### Patient group analysis

As previously mentioned, SEEG recordings involve multiple electrodes, typically ranging from 12 to 18, with each electrode containing up to 15 contacts (typically 10). Some contacts might record signals from the same IED ‘generators’. Consequently, bipolar channels with the highest concentration of IEDs are primary generators, and other channels may exhibit propagated IEDs.^21^ Moreover, a growing body of evidence suggests that high-frequency CS can lead not only to local but also remote activations.^22^ In essence, high-frequency CS delivered via SEEG primarily influences the targeted area, yet it can also impact neighboring and interconnected brain regions. In this context, contacts within the SOZ are thought to be more fragile, making them highly sensitive to minor perturbations.^23^ Thus, small cortical stimulations elsewhere might influence SOZ due to this heightened sensitivity.

As a result, we detected IEDs not only in the channels subjected to stimulation but also in all other non-stimulated contacts. In the next step, we calculated the IED rate (number of IED per min), and the IED rate change (IED rate (after stimulation) – IED rate (before stimulation)). To maintain consistency across all patient datasets, we normalized the IED rate changes by the maximum IED rate change observed within each patient, resulting in values between 0 and 1 (IED rate change/ maximum IED rate change). The “maximum IED rate change” refers to the highest IED rate change at any contact within that patient. We also built the epileptogenicity map of the patients based on the total number of IEDs in each contact during stimulation session.

### Effect of CS in non-stimulated contacts

We quantified IED rate changes at two levels: first, in channels directly subjected to stimulation to evaluate local effects, and second, in non-stimulated channels to assess whether cortical stimulation influences remote sites and whether such effects could aid in SOZ localization. For non-stimulated channels, we focused on the highest IED rate change observed at each contact. This approach allowed us to capture the most significant effect of stimulation while excluding minor or repeated fluctuations that could confound analyses or reduce classifier performance. Normalization was applied as described above, ensuring comparability across contacts and patients. ROC analyses were then performed to evaluate the predictive value of IED rate changes in both stimulated and non-stimulated contacts. This procedure enabled a clear assessment of how high-frequency cortical stimulation can propagate through interconnected brain networks and influence epileptogenic activity beyond the directly stimulated site.

### Statistical analysis

The delineation of SOZ and non-SOZ regions was achieved by considering various examination results and a comprehensive clinical interpretation (interictal and ictal SEEG recordings). These identified areas were subsequently validated in the selected patient cohort (as detailed in **Table 1**), where individuals achieved seizure freedom post-treatment. Consequently, each contact within the intracranial electrode array was categorized based on this classification. Our analytical approach involved conducting two distinct analyses, one focusing on contacts directly subjected to stimulation and the other on those not stimulated. To assess differences between the SOZ and non-SOZ groups, we applied a linear mixed-effects model (to account for repeated measures within patients, with SOZ status (SOZ vs non-SOZ) as a fixed effect and patient as a random effect (*p < 0.001*)). Furthermore, we utilized the receiver operating characteristic (ROC) curve methodology to determine an optimal threshold value for the bimodal distribution, facilitating the classification of SOZ and non-SOZ regions. To improve the specificity and sensitivity of the classification in areas not stimulated, we focused on the highest change evoked at each contact following stimulation, rather than examining all recordings in these regions.

### Labeling contacts

Initially, SOZ and non-SOZ were determined through examination results and supplementary clinical interpretation (interictal and ictal SEEG recordings) done by two epileptologists. These identifications were confirmed by the seizure-free outcomes of the patients after undergoing surgery or ablation. In cases where a stimulation pair included both SOZ and non-SOZ contacts, the pair was classified as SOZ. This approach was chosen to reflect the fact that stimulation involving any SOZ contact can directly engage the epileptogenic network.

## Results

### Example patient

We first discuss the results of a right-hand dominant patient diagnosed with focal epilepsy suspected to be of right temporal lobe origin. The patient was implanted with six pairs of electrodes targeting the left and right amygdala, insula, anterior hippocampus, posterior hippocampus, orbital frontal, and posterior cingulate (**Fig. 4A**). Delivering stimulation over bipolar channels can cause different events depending on whether they are in the epileptogenic zone or the propagation zone. These events can include afterdischarges, electrical events such as subclinical EEG seizures, electroclinical events such as stimulation-induced seizures, and no stimulation-induced events (normal).^24^ The patient underwent 12 stimulation trials across both hemispheres, starting with the least epileptogenic channels and progressing to the most epileptogenic ones (see supplementary **Table 2**). The degree of epileptogenicity was determined during pre-stimulation monitoring by quantifying the ictal discharges observed in those channels.

**Figure 4.**
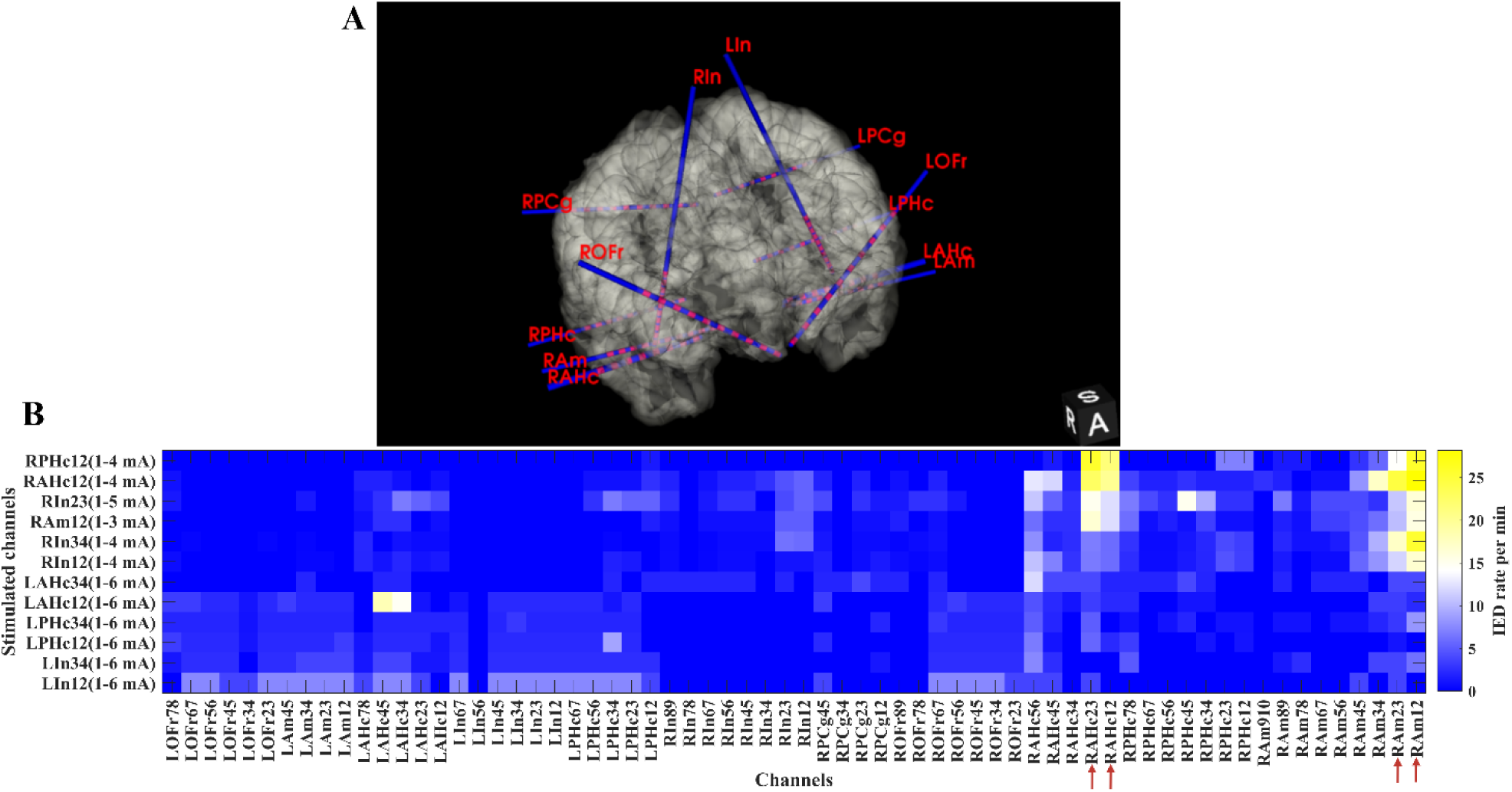
Epileptic activity following stimulation. **(A)** 3D view of depth electrode locations for a representative patient. Depth electrodes sampled the following regions: Left hemisphere — Left Amygdala (LAm), Left Anterior Hippocampus (LAHc), Left Posterior Hippocampus (LPHc), Left Insula (LIn), Left Orbital Frontal cortex (LOFr), Left Posterior Cingulate (LPCg); Right hemisphere — Right Amygdala (RAm), Right Anterior Hippocampus (RAHc), Right Posterior Hippocampus (RPHc), Right Insula (RIn), Right Orbital Frontal cortex (ROFr), Right Posterior Cingulate (RPCg). **(B)** IED generation pattern over all channels after each stimulation trial. The x-axis lists all channels after applying a bipolar montage (e.g., RAm12 denotes the bipolar derivation between contacts RAm-1 and RAm-2). Noisy channels were excluded. The y-axis indicates the stimulated bipolar channels and the range of delivered current through them (currents varied across trials and channels; e.g., 1–6 mA). SOZ is associated with highest evoked IEDs which are associated with RAm channels 1, 2, 3 and RAHc channels 1, 2, 3 (marked by red arrows). IED: interictal epileptiform discharge. Panels A and B were generated in 3D Slicer and MATLAB and combined using Biorender. (Created in BioRender. Ahmadi, A. (2026) https://BioRender.com/y01fzx7)

The bipolar stimulation trials were conducted over the following channels: left insula channels 1 and 2, left insula channels 3 and 4, left posterior hippocampus channels 1 and 2, left posterior hippocampus channels 3 and 4, left anterior hippocampus channels 1 and 2, left anterior hippocampus channels 3 and 4, right insula channels 1 and 2, right insula channels 3 and 4, right amygdala channels 1 and 2, right insula channels 2 and 3, right anterior hippocampus channels 1 and 2 and finally, right posterior hippocampus channels 1 and 2.

As shown in supplementary **Table 2**, stimulating the epileptogenic and SOZ identified during intracranial monitoring led to the emergence of afterdischarges and CS-evoked seizures. The maximum current delivered was adjusted based on the responses observed after each stimulation trial. Considering all the factors discussed in IED detection section, the rate of evoked IEDs was computed following each stimulation trial, with its pattern of occurrence across all channels depicted in **Fig. 4B**.

Stimulating the SOZ and adjacent areas resulted in a marked increase in IED rates ( **Fig. 4B**). This elevated IED rate is also evident when adjacent areas to the SOZ are stimulated, suggesting that the enhanced excitability of the SOZ extends to neighboring regions. By utilizing the cumulative number of IEDs recorded after each stimulation trial, we constructed an epileptogenicity map of the brain (with the exclusion of noisy channels) (**Fig. 5A**). Stimulation of regions that produced a lower number of IEDs did not trigger seizures. In contrast, stimulation of areas with the highest IED counts during the session induced seizures and afterdischarges. In this patient, the right amygdala and hippocampus exhibited significant epileptogenicity, producing afterdischarges and CS-evoked seizures. These findings indicate that the epileptogenic network involves the right mesial temporal region, including the amygdala and hippocampus, which also generated the highest number of IEDs during the stimulation session. The epileptogenicity map of the patient can also be constructed using patient’s MRI scans (**Fig. 5B**).

**Figure 5.**
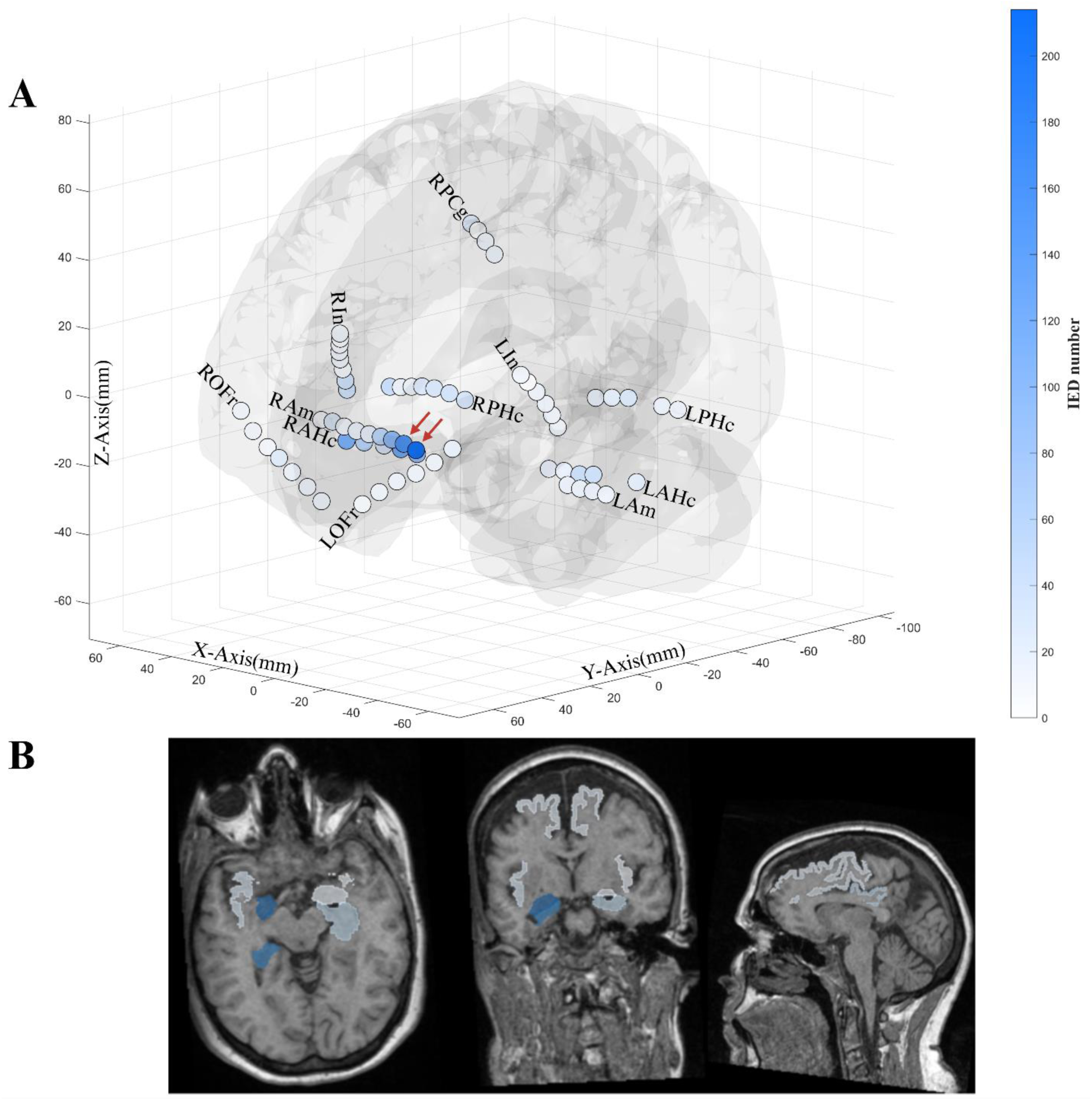
Epileptogenicity maps. **(A)** 3D Epileptogenicity map based on the total number of IEDs. Electrode contacts are color-coded according to the number of IEDs (dark blue colors represent higher IED number (marked by red arrows). IED: interictal epileptiform discharges; RPHc: right posterior hippocampus; RAHc: right anterior hippocampus; RIn: right insula; RAm: right amygdala; RPCg: right posterior cingulate; ROFr: right orbitofrontal; LAHc: left anterior hippocampus; LPHc: left posterior hippocampus; LIn: left insula; LAm: left amygdala; LOFr: left orbitofrontal. **(B)** Epileptogenicity map on patient’s MRI scan. The solid segments indicate the areas monitored with depth electrodes (right hippocampus, right insula, right amygdala, right orbitofrontal, right posterior cingulate, left hippocampus, left insula, left amygdala, left orbitofrontal). The dark blue regions correspond to the areas that evoked higher IEDs following stimulation. The color for each zone corresponds to the contact in that zone that generated the highest IED number post stimulation. From left to right: the axial, coronal, and sagittal views. MRI: Magnetic Resonance Imaging. Panels A and B were generated in MATLAB and 3D Slicer and combined using Biorender. (Created in BioRender. Ahmadi, A. (2025) https://BioRender.com/wp3a6w5)

### Patient group analyses

A total of 328 electrodes were implanted, yielding 3078 contacts. Among these, 538 contacts were classified as SOZ and 1847 contacts as non-SOZ. The remaining contacts were excluded from the analyses because they were either noisy or located in white matter. To maintain consistency in results across all 30 patients, we calculated the normalized absolute change in IED rates for each contact point. This method allowed us to standardize the data, ensuring reliable comparisons of IED changes across patients and stimulation sites. As previously mentioned, these patients were seizure-free post-surgery, enabling us to delineate SOZ and non-SOZs based on clinical assessments and surgical outcomes. The changes in IEDs’ activity at the stimulation sites are shown in **Fig. 6**. The analysis demonstrates a statistically significant difference in CS-evoked IEDs between SOZ and non-SOZ regions (linear mixed effect model, *p < 0.001, β= -0.40, R^2^= 0.64*).

**Figure 6.**
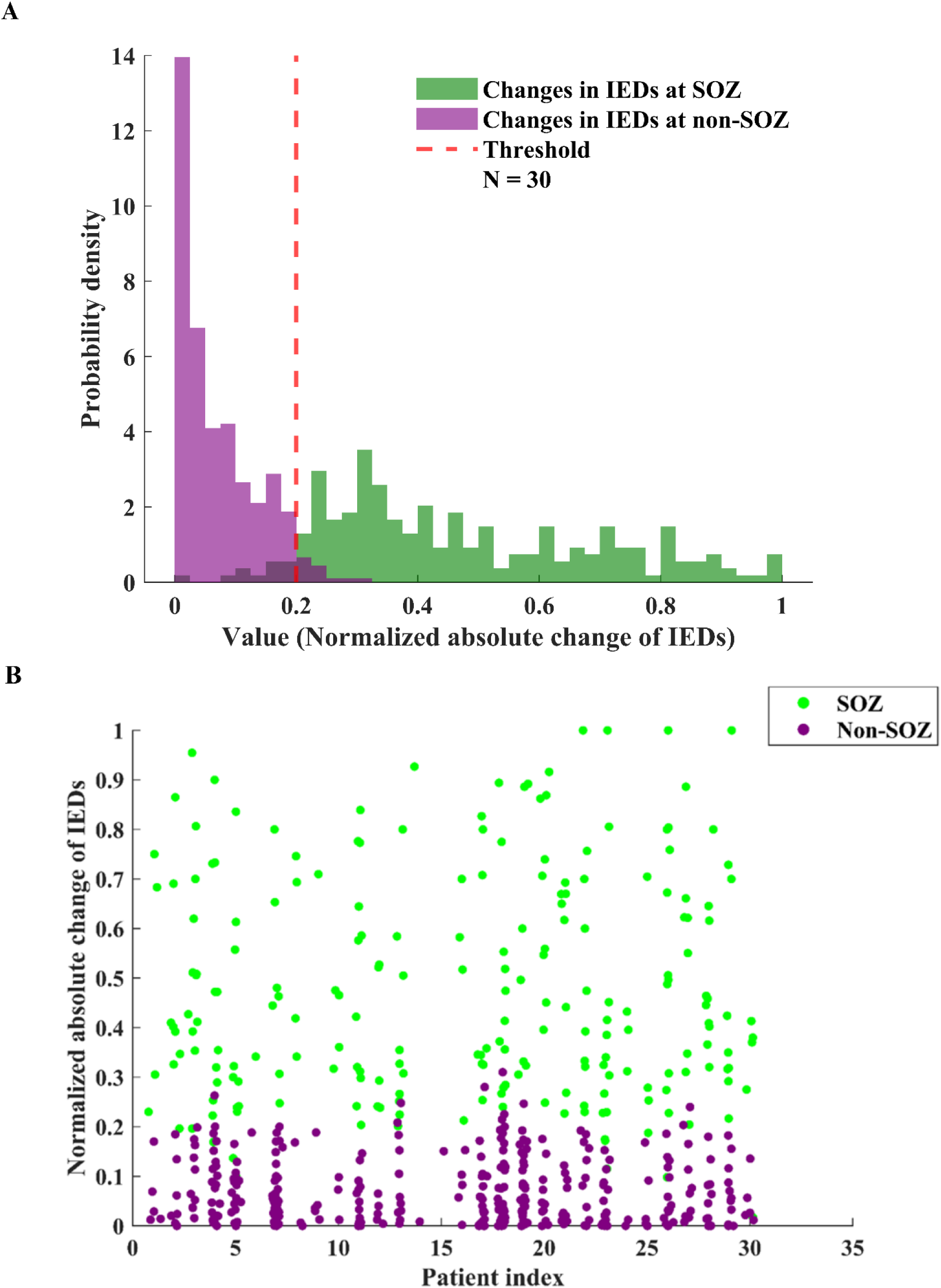
The effect of stimulation at the SOZ and non-SOZ (site of stimulation). **(A)** Comparison of normalized IED changes between SOZ and Non-SOZ. The x-axis shows the normalized absolute change in IED rate, which is a measure of change relative to a baseline (observe that the rates have been normalized to the highest rate for each patient). The y-axis shows the probability density of the recordings per value. Green bars represent changes in IEDs at the SOZ after SOZ stimulation and the purple bars represent changes in IEDs at non-SOZ regions after non-SOZ stimulation. Asignificant difference between the two groups (linear mixed effect model, *p-value <0.001, β= -0.40, R^2^= 0.64*). *Threshold value: 0.2* (ROC). *Specificity: 0.97*. *Sensitivity: 0.94*. *AUC: 0.98*. *N= 30 patients*. IED: Interictal epileptiform discharges; SOZ: Seizure onset zone; non-SOZ: non-seizure onset zone. N: Number of patients; ROC: receiver operating characteristic; AUC: Area under the curve. **(B)** Scatter plot showing IED rate changes per patient. The x-axis shows patients’ index. The y-axis shows the normalized absolute change in IED rate, which is a measure of change relative to a baseline (observe the rates have been normalized to the highest rate for each patient). Each point represents individual contact, with slight horizontal jitter added for visualization clarity. Green points show stimulated contacts in SOZ, and purple points show stimulated contacts in non-SOZ. A significant difference between the two groups (linear mixed effect model, *p-value <0.001, β= -0.40, R^2^= 0.64, Number of patients= 30*). IED: Interictal epileptiform discharges; SOZ: Seizure onset zone; non-SOZ: non-seizure onset zone. Panels A and B were generated in MATLAB and combined using Biorender. (Created in BioRender. Ahmadi, A. (2026) https://BioRender.com/n20aw7n)

To develop our quantitative method for SOZ identification, we employed a ROC curve to determine an optimal threshold value for a bimodal classifier. The red dashed line indicates a threshold value, which is used to classify recordings into SOZ or non-SOZ. The high population density of the purple bar indicates that a significant number of non-SOZ regions experienced minimal changes in IED rate activity following stimulation. A *specificity* of *0.97* indicates a high true negative rate and a *sensitivity* of *0.94* indicates a high true positive rate. *Area under the curve* of *0.98*, suggests high discrimination between SOZ and non-SOZ based on IED rate change. This suggests that non-SOZ areas typically have less excitability or response to stimulation compared to SOZ regions, which aligns with the expected behavior where SOZ regions are more responsive to stimulation due to their epileptogenic nature (see Supplementary **Fig. 4** for ROC analysis).

### Effect of CS in non-stimulated contacts

The second component of the analysis focused on contacts other than the stimulated ones. Specifically, we examined how stimulation of a given pair of contacts influenced IED activity across all other non-stimulated contacts. The objective was to determine whether stimulation produced measurable changes in IED rates at non-stimulated regions and to assess whether these effects were related to the localization of the SOZ. We observed a significant difference in IED rate changes between SOZ and non-SOZ regions (linear mixed effect model, *p < 0.001, β= -0.09, R^2^= 0.15*). Consistent with our approach at stimulation sites, we used ROC curve analysis to determine an optimal threshold for SOZ identification. In this context, a *threshold value* of *0.05* was established, with a *specificity* of *0.73*, *sensitivity* of *0.57*, and an *AUC* of *0.71* (see Supplementary **Fig. 1**, **Fig. 2** and **Fig. 3**). To improve the specificity and sensitivity of the bimodal classification in this case, we focused on the highest IED change evoked at each contact following stimulation, rather than examining all recordings in non-stimulated regions. **Fig 7** shows the highest IED change evoked at each contact in the areas that were not stimulated. The results indicate an increase in the rate of IEDs at the SOZ, even in regions that were not directly stimulated (linear mixed effect model, *p-value <0.001, β= -0.44, R^2^= 0.67*). This demonstrates that the effect of stimulation extends beyond the immediate site of stimulation.

**Figure 7.**
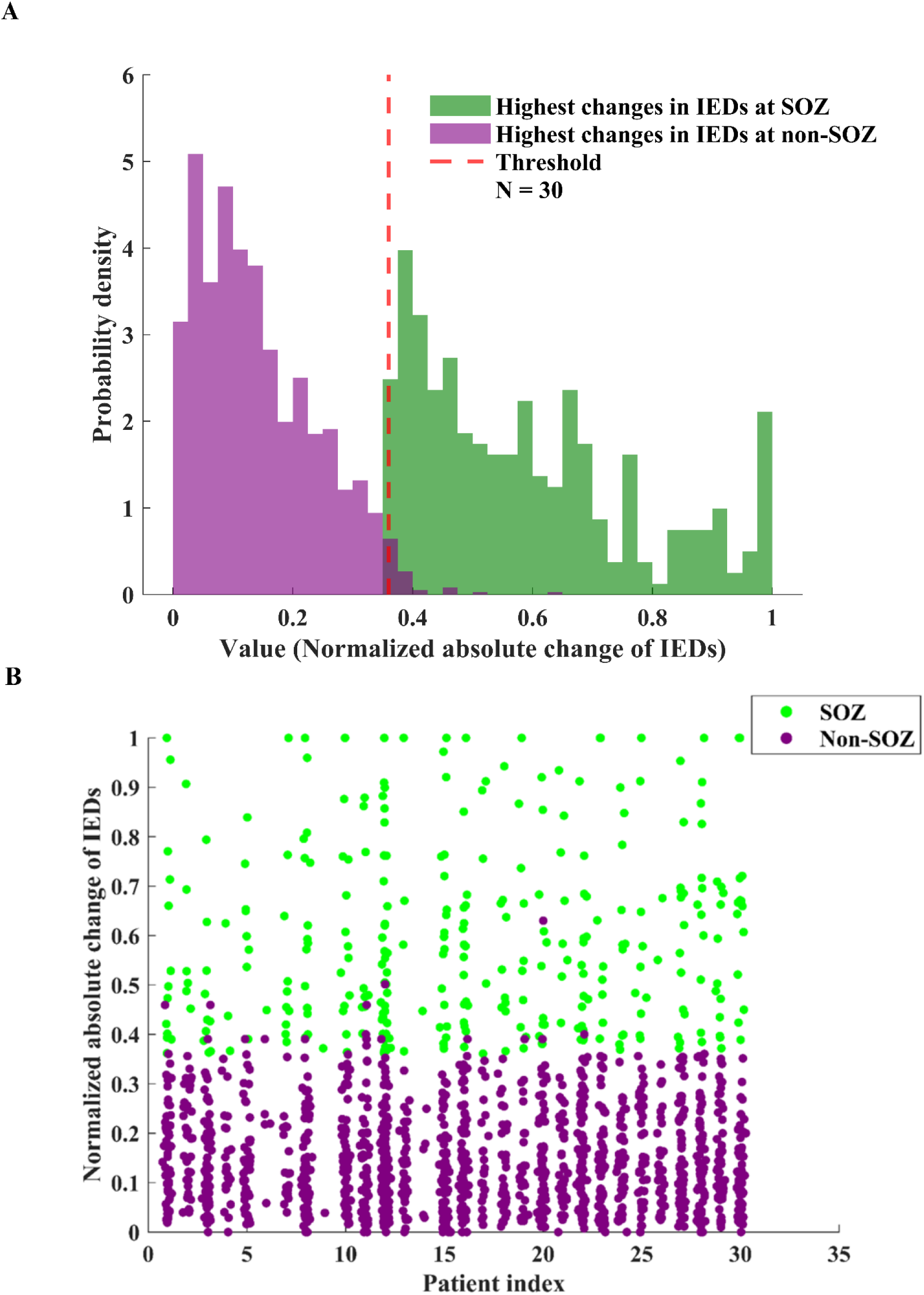
The effect of stimulation at non-stimulated areas (highest changes recorded at non-stimulated areas). **(A)** Comparison of normalized IED changes between SOZ and Non-SOZ. The x-axis shows the normalized absolute change in IED rate, which is a measure of change relative to a baseline (observe the rates have been normalized to the highest rate for each patient). The y-axis shows the probability density of the recordings per value. Green bars represent highest changes in IEDs rate (recorded at non-stimulated areas) at the SOZ and the purple bars represent highest changes in IEDs at non-SOZ after stimulation. A significant difference between the two groups (linear mixed effect model, *p-value <0.001, β= -0.44, R^2^= 0.67*). *Threshold value: 0.36 (ROC). Specificity: 0.98. Sensitivity: 1. AUC: 0.99*. *N= 30 patients*. IED: Interictal epileptiform discharges; SOZ: Seizure onset zone; non-SOZ: non-seizure onset zone. N: Number of patients; ROC: receiver operating characteristic; AUC: Area under the curve. **(B)** Scatter plot showing IED rate changes per patient. The x-axis shows patients’ index. The y-axis shows the normalized absolute change in IED rate, which is a measure of change relative to a baseline (observe the rates have been normalized to the highest rate for each patient). Each point represents individual contact, with slight horizontal jitter added for visualization clarity. Green points represent highest changes in IEDs rate recorded from each contact (at non-stimulated areas) at the SOZ and the purple points represent highest changes in IEDs at non-SOZ after stimulation. Asignificant difference between the two groups (linear mixed effect model, *p-value <0.001, β= -0.44, R^2^= 0.67, Number of patients= 30*). IED: Interictal epileptiform discharges; SOZ: Seizure onset zone; non-SOZ: non-seizure onset zone. Panels A and B were generated in MATLAB and combined using Biorender. (Created in BioRender. Ahmadi, A. (2026) https://BioRender.com/5wrc0ne)

## Discussion

In the current study, we examine whether analyses of non-ictal events, the IED evoked by CS can aid the localization of the SOZ in patients with DRE evaluated for surgical treatment. We quantified the rate of IEDs in different regions before and after the CS procedure and quantified the rate of IED within and outside the SOZ determined by standard procedures (clinical and analyses of spontaneous ictal activity) and found that CS selectively increases the rate of IED within the SOZ. An ROC analyses indicated it is possible to localize the electrodes within the SOZ by examining the rate of stimulation evoked IEDs and compare it with the non-SOZ rates.

Our study revealed a significant change in IEDs at the SOZ following CS, even in regions not directly stimulated, indicating that CS-induced IED can serve as a reliable biomarker for identifying epileptogenic zones. Our procedure can also offer valuable insights into the underlying pathophysiology of epileptogenic networks in patients with DRE. Quantitative analysis of CS-evoked IED changes at stimulated and non-stimulated areas revealed a distinction between SOZ and non-SOZ regions. The latter can aid surgical interventions in DRE patients, ultimately leading to improved seizure management and quality of life.

### Candidate mechanisms of IED increase during CS

The observed increase in the rate of IEDs following CS can be attributed to several underlying mechanisms. First, stimulation induces depolarization of neurons in the vicinity of the stimulation sites, leading to an increased likelihood of synchronous firing. This phenomenon, known as depolarization block, results from the accumulation of sodium ions within neurons, causing a transient suppression of neuronal activity followed by a period of hyperexcitability.^25^ Second, CS may alter the balance of excitatory and inhibitory neurotransmission, potentially leading to an overall increase in neuronal excitability within the stimulated region.^26^ The differential response of channels in SOZ and non-SOZ to CS underscores the underlying pathophysiological distinctions between these regions. Within the SOZ, the neural circuitry is characterized by a lower threshold for synchronized neuronal firing, likely due to an imbalance between excitatory and inhibitory inputs.^2^ CS may disrupt this delicate balance, leading to an increase in IED frequency. Conversely, in non-epileptogenic zones, the inhibitory mechanisms are more effective in suppressing abnormal neuronal firing, resulting in a comparatively muted response to CS.^27^

The study’s findings align with the existing body of literature that highlights the intricate interplay between stimulation and neural network activation.^27–33^ It is well-established that IEDs arise from the aberrant synchronization of neuronal populations, often driven by local excitatory-inhibitory imbalances. CS, acting as an external perturbation, further modulates this delicate network, potentially exacerbating the generation and propagation of IEDs. The observed increase in IEDs following CS in the channels where seizures were triggered provides empirical evidence of this phenomenon.

Interestingly, neural stimulation techniques have been utilized to inhibit epileptic seizures. Responsive neurostimulation uses frequencies of 200 Hz to inhibit ictal discharges.^34^ Although 200 Hz is the typical initial programming setting, lower frequencies (as low as 5 Hz) are also used in clinical practice when higher frequencies are suboptimal or to minimize stimulation-related side effects, and stimulation durations can range from 100 ms to several seconds. Additionally, in open-loop anterior thalamic stimulation for epilepsy, the default frequency is typically around 145 Hz. Although the results are variable across patients and the mechanisms of action of RNS remain largely unclear, it has been proposed that high-frequency stimulation at the SOZ reduces the synchrony in epileptogenic networks and therefore the probability of seizures.^35^ Interestingly, in animal models, it has been shown that low-frequency stimulation (1-8 Hz) using optogenetics suppresses epileptogenesis.^36^ This variety of results seems to indicate that it may be crucial to consider the parameters of the CS, including the applied frequency, when designing diagnostic or therapeutic interventions. Clarifying these issues may require studies involving large sample sizes as well as animal models in which precise manipulations of neurons and networks are more feasible.

### Limitations and Future Directions

Standardized stimulation parameters and protocols at our institution ensured consistency across patients, enhancing the reliability and reproducibility of our findings. However, our study is not without limitations. The results may have limited generalizability due to the study’s single-site design and the relatively small number of patients included. Future studies with larger cohorts and multi-center collaborations are warranted to validate the findings and further elucidate the role of stimulation on IEDs in SOZ localization. A long-term follow-up study is necessary to assess the durability of surgical outcomes and the risk of spontaneous seizure recurrence.

Additionally, the number of electrodes implanted, and the implantation areas varied across patients, as did the stimulation areas. This inconsistency prevented us from performing a structure-based analysis applicable to all patients. Furthermore, the pathology of epilepsy can influence biomarkers in intracranial electroencephalography data. The specific characteristics and patterns of activity detected in SEEG can differ based on the underlying pathology causing the epilepsy. Our cohort included patients with diverse pathologies and epilepsy types. Future studies with larger sample sizes could focus on specific pathology and epilepsy type to validate and extend our results.

Another potential limitation of this study is that the distance between stimulated and recording contacts was not explicitly modeled. Because electrode implantation schemes and stimulation pairs varied across patients, a systematic quantification of distance effects was not feasible with the current dataset. Nevertheless, both SOZ and non-SOZ regions included contacts located at various distances from the stimulation sites, which likely mitigates potential distance-related bias.

Moreover, applying this analysis to patients who did not achieve seizure freedom following intervention would be valuable to assess whether the seizure onset zone identified using this approach extends beyond the resected area. However, such an analysis was beyond the scope of the present study. As our cohort grows, we plan to include a larger sample of both seizure-free and non–seizure-free patients to enable internal validation of the current findings.

### Significance

Our results, derived from a single session of stimulation, revealed information comparable to that obtained from intracranial monitoring conducted over several days, as well as from other established modalities for SOZ identification. This finding highlights the critical importance of data gleaned from stimulation sessions. Such efficiency not only reduces the time required for effective diagnosis but also minimizes the risks and discomforts associated with prolonged invasive monitoring.

The ability to rely on stimulation-based information is particularly vital in scenarios where ictal activity does not occur during presurgical monitoring. Additionally, it becomes crucial when prolonged patient hospitalization is not feasible due to infection risks or other medical complications. In such cases, the efficiency of single-session CS in providing critical diagnostic information ensures that patients receive timely and accurate evaluations, avoiding extended and potentially hazardous monitoring periods.

In certain instances, it is challenging to evoke patients’ typical seizures due to specific limitations of the patient’s condition. Consequently, stimulation may need to be limited to language mapping while avoiding areas that might evoke seizures. Therefore, monitoring the effect of CS on interictal phenomena can be critical.

## Conclusions

In summary, this research illustrated the excitatory effects of high-frequency intracortical stimulation at the SOZ, as evidenced by an increase in the rate of IEDs, suggesting that stimulation-induced IEDs can be used as a biomarker to identify the SOZ. Our study underscores the transformative potential of high-frequency CS in the management of DRE, leading to personalized and tailored treatment strategies and subsequently improved patient outcomes. By leveraging advanced neurophysiological techniques and computational approaches, we can unravel the complex dynamics of epileptogenic networks and develop targeted interventions tailored to each patient’s unique pathophysiology.

## Data Availability

All data produced in the present study are available upon reasonable request to the authors.

