## Supplementary material for "50 Hz cortical stimulation increases interictal epileptiform discharges at the seizure onset zone"

**Supplementary Table 1 Stimulation parameters**

| Parameters for High Frequency | |
|---|---|
| Stimulation Mode | Bipolar |
| Stimulation Frequency | 50 Hz |
| Current | 1–8 mA |
| Stimulation Time | 5 s |
| Interval InterStimulations | ≥10 s |
| Pulse Width | 300 μs |

**Supplementary Table 2 Stimulation trials**

| Trial | Channels | Current | Event |
|---|---|---|---|
| 1 | LIn 1-2 | 1-6 mA | Normal |
| 2 | LIn 3-4 | 1-6 mA | Normal |
| 3 | LPHc 1-2 | 1-6 mA | Normal |
| 4 | LPHc 3-4 | 1-6 mA | Normal |
| 5 | LAHc 1-2 | 1-6 mA | Afterdischarges |
| 6 | LAHc 3-4 | 1-6 mA | Normal |
| 7 | RIn 1-2 | 1-4 mA | Normal |
| 8 | RIn 3-4 | 1-4 mA | Afterdischarges |
| 9 | RAm 1-2 | 1-3 mA | Seizure |
| 10 | RIn 2-3 | 1-5 mA | Afterdischarges |
| 11 | RAHc 1-2 | 1-4 mA | Seizure |
| 12 | RPHc 1-2 | 1-4 mA | Afterdischarges |

LIn: Left insula; LPHc: left posterior hippocampus; LAHc: left anterior hippocampus; RIn: Right insula; RAm: right amygdala; RAHc: right anterior hippocampus; RPHc: right posterior hippocampus.

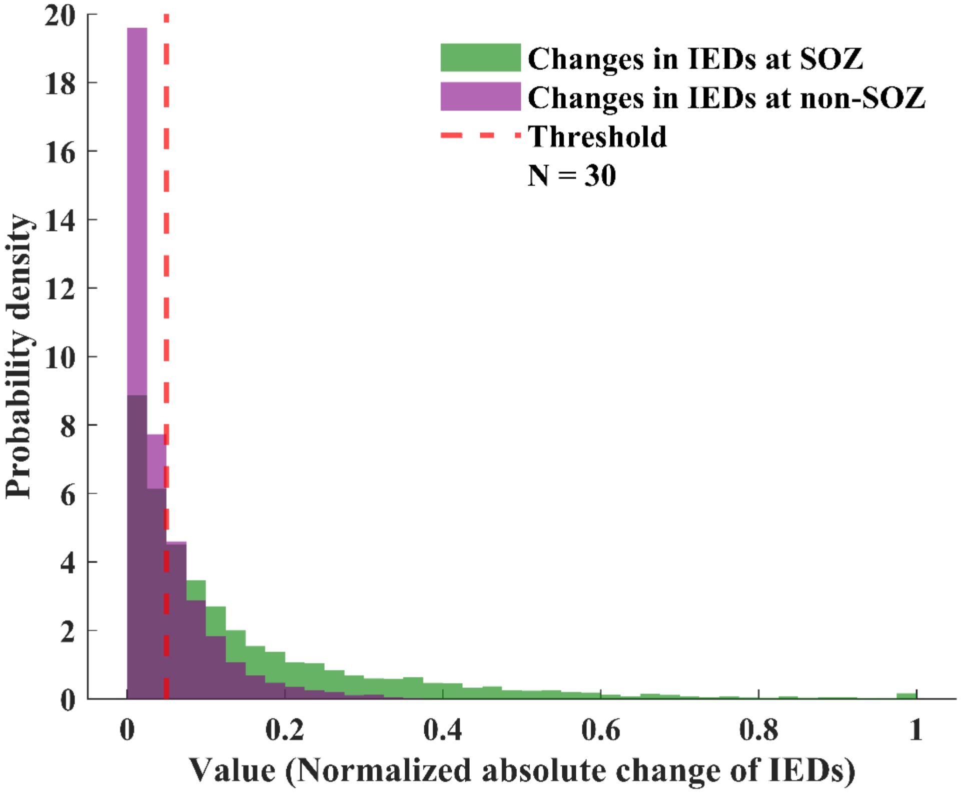


**Supplementary Figure 1 The effect of stimulation at non-stimulated areas.** The x-axis shows the normalized absolute change in IED rate, which is a measure of change relative to a baseline (observe the rates have been normalized to the highest rate for each patient). The y-axis shows the probability density of the recordings per value. Green bars represent changes in IEDs rate (recorded at non-stimulated areas) at the SOZ and the purple bars represent changes in IEDs at non-SOZ after stimulation. A significant difference between the two groups (linear mixed effect model, *p-value <0.001*). *Threshold value*: *0.05* (ROC). *Specificity*: *0.73*. *Sensitivity*: *0.57*. *AUC*: *0.71*. *N= 30 patients*. IED: Interictal epileptiform discharges; SOZ: Seizure onset zone; non-SOZ: non-seizure onset zone. N: Number of patients; ROC: receiver operating characteristic; AUC: Area under the curve.

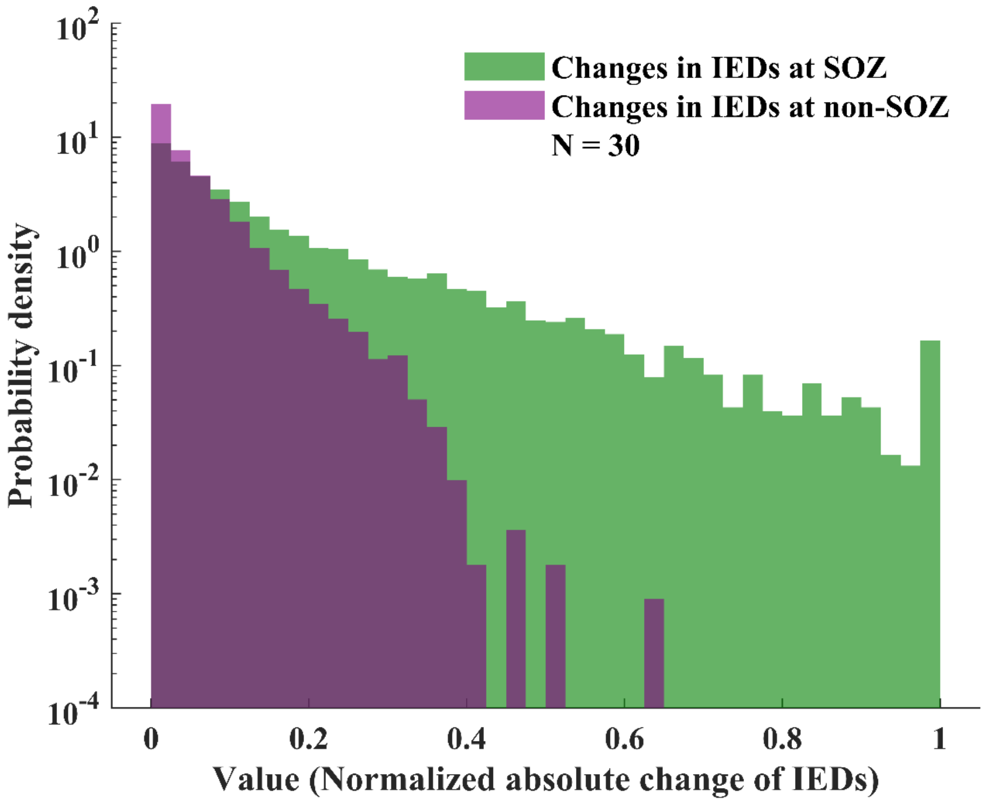


**Supplementary Figure 2 The logarithmic presentation of Figure 1**. The x-axis shows the normalized absolute change in IED rate, which is a measure of change relative to a baseline (observe the rates have been normalized to the highest rate for each patient). The y-axis shows the probability density of the recordings per value (logarithmic values). Green bars represent changes in IEDs rate (recorded at non-stimulated areas) at the SOZ and the purple bars represent changes in IEDs at non-SOZ after stimulation. A significant difference between the two groups (linear mixed effect model, *p-value <0.001*) *N=30 patients*. IED: Interictal epileptiform discharges; SOZ: Seizure onset zone; non-SOZ: non-seizure onset zone. N: Number of patients.

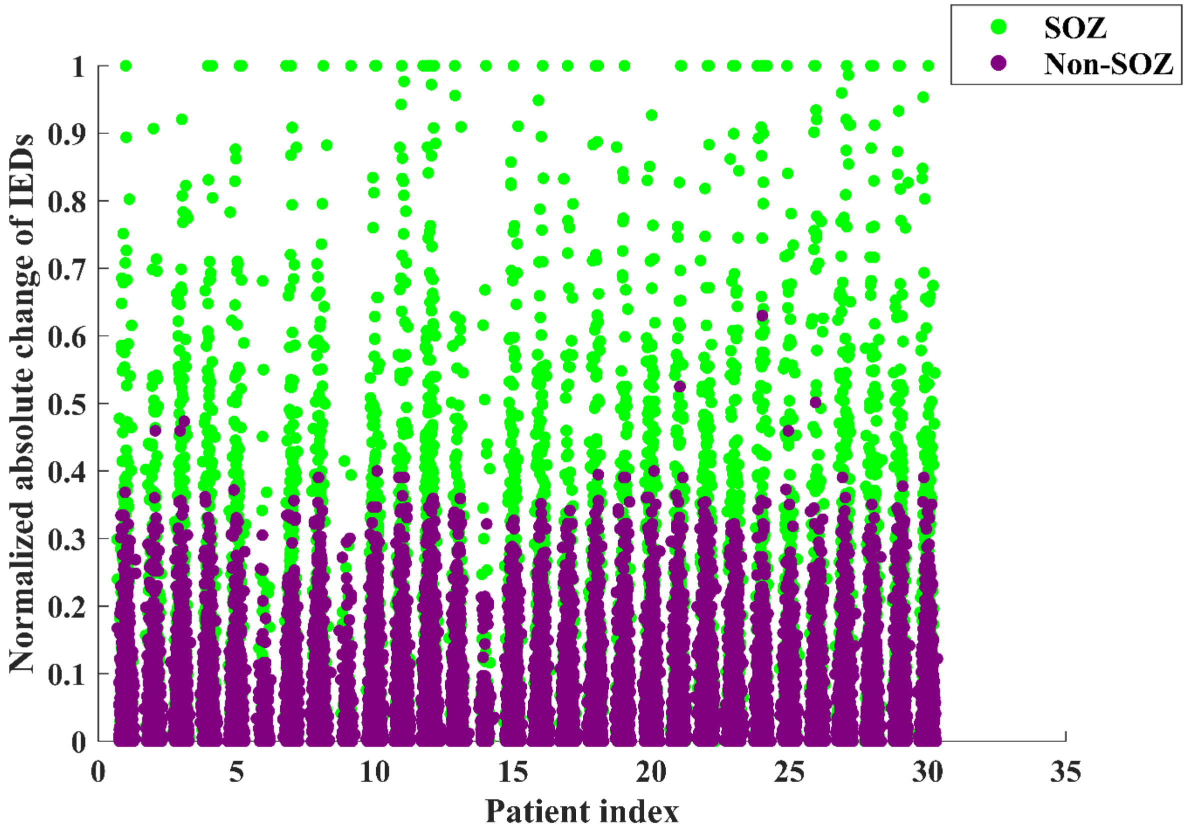


**Supplementary Figure 3 Scatter plot showing IED rate changes per patient (at non-stimulated sites).** The x-axis shows patients' index. The y-axis shows the normalized absolute change in IED rate, which is a measure of change relative to a baseline (observe the rates have been normalized to the highest rate for each patient). Each point represents individual contact, with slight horizontal jitter added for visualization clarity. Green points represent changes in IEDs rate (recorded at non-stimulated areas) at the SOZ and the purple points represent changes in IEDs at non-SOZ after stimulation. A significant difference between the two groups (linear mixed effect model, *p-value* $<0.001$, $\beta = -0.09$, $R^2 = 0.15$, *Number of patients* $= 30$). IED: Interictal epileptiform discharges; SOZ: Seizure onset zone; non-SOZ: non-seizure onset zone.

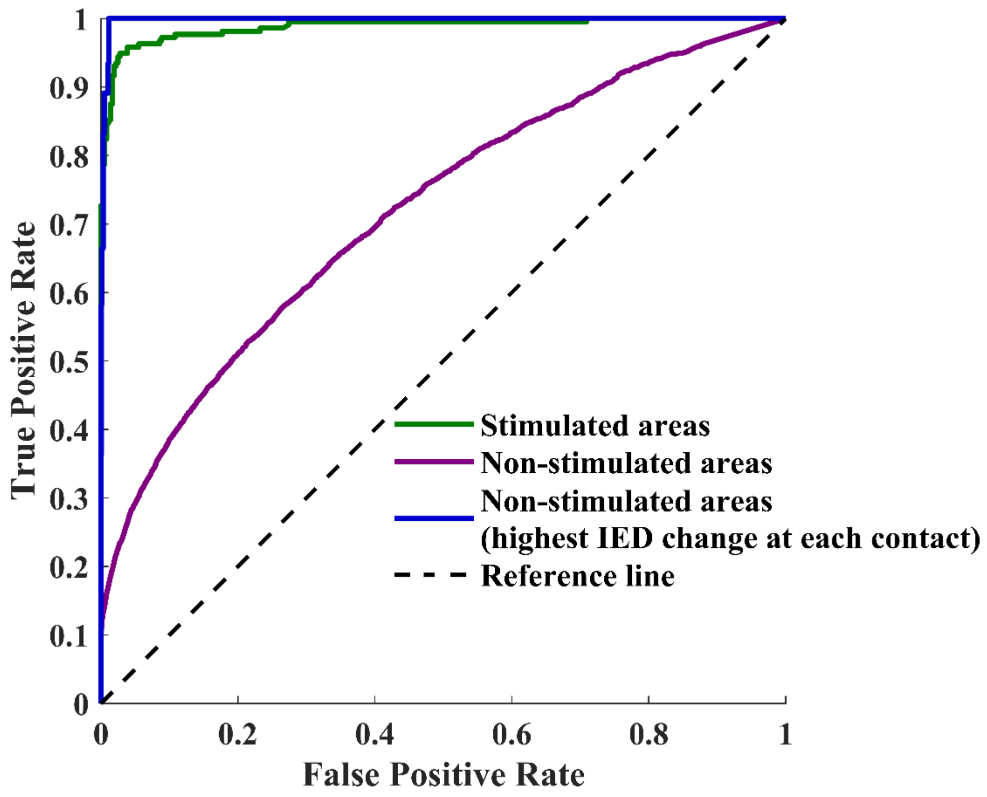


**Supplementary Figure 4 ROC analysis illustrates the effect of cortical stimulation on IEDs.** The green line represents the effect of stimulation at stimulated areas (*threshold: 0.20, specificity: 0.97, sensitivity: 0.94, AUC: 0.98, 95% CI = [0.979 - 0.995], bootstrap test, p-value<0.001*). The purple line represents the effect of stimulation at non-stimulated areas (*threshold: 0.05, specificity: 0.73, sensitivity: 0.57, AUC: 0.71, 95% CI = [0.714 - 0.724], bootstrap test, p-value<0.001*). The blue line represents the effect of stimulation at non-stimulated areas considering only the highest IED changes at each contact (*threshold: 0.36, specificity: 0.98, sensitivity: 1.0, AUC: 0.99, 95% CI = [0.996 - 0.999], bootstrap test, p-value<0.001*). The black dashed line represents the reference line at chance level (*AUC: 0.5*). IED: Interictal epileptiform discharges; ROC: receiver operating characteristic; AUC: area under the curve; CI: Confidence Interval.
